# Should contact bans be lifted in Germany? A quantitative prediction of its effects

**DOI:** 10.1101/2020.04.10.20060301

**Authors:** Jean Roch Donsimoni, René Glawion, Bodo Plachter, Constantin Weiser, Klaus Wälde

## Abstract

Many countries consider the lifting of restrictions of social contacts (RSC). We quantify the effects of RSC for Germany. We initially employ a purely statistical approach to predicting prevalence of COVID19 if RSC were upheld after April 20. We employ these findings and feed them into our theoretical model. We find that the peak of the number of sick individuals would be reached already mid April. The number of sick individuals would fall below 1,000 at the beginning of July. When restrictions are lifted completely on April 20, the number of sick should rise quickly again from around April 27. A balance between economic and individual costs of RSC and public health objectives consists in lifting RSC for activities that have high economic benefits but low health costs. In the absence of large-scale representative testing of CoV-2 infections, these activities can most easily be identified if federal states of Germany adopted exit strategies that *differ* across states.

## 1 Introduction

Authorities in most countries have imposed restrictions on social contacts (RSC in what follows) in various forms. They include contact bans outside the household, shut down of schools and closing of small businesses. Many countries are facing the question of how long RSC should last. We take the example of Germany and quantify both their current effects and their effects in the long run in case they are maintained. We also quantify the effect of a complete lift of RSC.

We find that neither a permanent RSC nor a complete lift is desirable. A permanent RSC would yield an epidemic in Germany that would lead to around 184,000 sick individuals only. The epidemic would not be over, however, as most individuals would still likely be susceptible to an infection. A permanent RSC would also not be economically sustainable. A complete lift is likely to yield a fast increase of the number of sick that would overstrain the public health system. This points towards the need to think about exit options which promise to keep infection rates stable. Exit strategies should be reversible and tested for, say, 4 weeks and differ across regions. This would allow authorities to understand their health and economic effects. Learning about policy measures appears essential in this global pandemic.

There is an exploding literature on COVID19 and its effects. A first survey is in Donsimoni et al. (2020), a broader overview is in Gros et al. (2020). We build our analysis on the model and projection presented in Donsimoni et al. (2020a).^2^ In contrast to this paper, we (i) provide a more precise calibration of the effect of no public health measures. The precision results from the availability of more observations. This is essential for quantifying the effects of lifting RSC. We (ii) can also quantify the effects of RSC in the present paper as sufficient data has become available since our earlier work. Our most recent observation now is from 7 April. Most importantly, due to the availability of enough observations, we can (iii) employ purely statistical methods to make a forecast *for the current RSC*. This allows us to work without assumptions about long-run infection and sickness rates. For judging the effect of a lift of RSC, we do need to return to long-run assumptions, however, as we need to work with the theoretical model developed in Donsimoni et al. (2020a) again.

Adamik et al. (2020) also quantitatively analyse the situation in Germany. They employ a microsimulation model which allows to better understand the effect of heterogeneity across households. They argue that reaching herd immunity without violating the capacity limit of the health care system is likely to fail. They do not explicitly analyse the effects of RSC and do not discuss the fit of their model to observed data. Dehning et al. (2020) estimate parameters of their model in a statistically very convincing way. They focus on constant transition rates for different RSC-regimes (but do allow for time-dependency to smooth between regimes). They make forecasts for a period of two to three weeks and use data up to 31 March.^3^ The analysis by Gros et al. (2020) also takes the economic costs of RSC into account. They do not provide forecasts. Promising future work could combine their economic cost approach with forecasts.

The structure of the paper is as follows. We first take a purely statistical perspective and describe the dynamics of the number of reported infected individuals over time. We employ both data from the Robert Koch Institute (RKI, 2020) and from Johns Hopkins University (JHU, 2020). We also provide a forecast of the number of reported sick individuals purely based on RKI observations and under the assumption that current RSC rules do not change. Section 3 presents the essentials of the model developed earlier in Donsimoni et al. (2020a). Our calibration is in section 4 and section 5 quantifies the effects of the current RSC and studies the effects of a complete exit. Section 6 concludes.

## 2 A first look at the data

### Descriptive statistics

There are two datasets for Germany that are used to describe prevalence of COVID19. The first is data from the Robert Koch Institute (RKI, 2020), the second data source is from Johns Hopkins University (JHU, 2020). In this section we employ both to see their relative strengths and merits.

When we look at figure 1, one might believe to identify a permanent break in growth rates end of March. Looking at the right picture gives the impression that the curve becomes flatter over time but there is a kink on 30 March: When looking at growth rates (crosses in left part of the figure), there is a permanent drop on this same 30 March.

**Figure 1.**
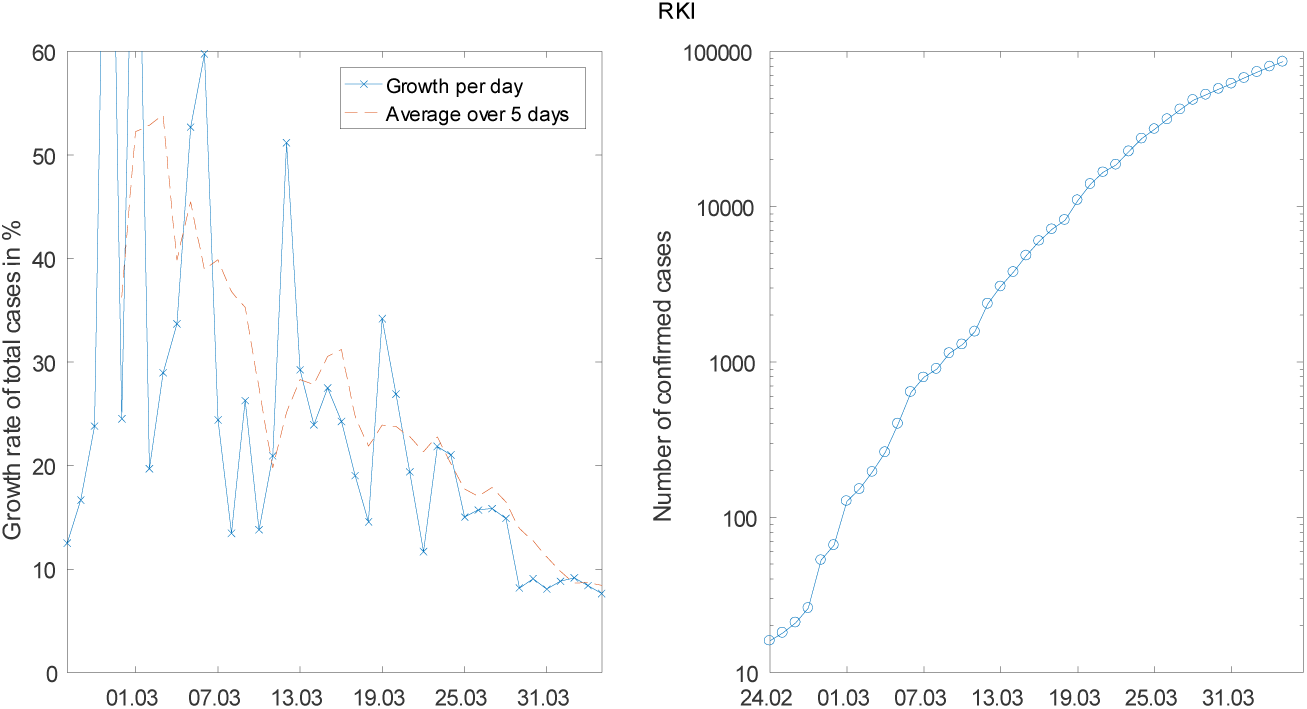
The daily growth rates (left) and the level of the number of sick (right) for RKI data (logarithmic scale)

When we look at Johns Hopkins data, we can identify two break points. The first is on 20 March. It can clearly be seen in the left part of the figure with the drop in daily growth rates and in the right part on 20 march. This is the drop that was also identified econometrically by Hartl et al. (2020). It is also clear from these two figures that there is another break on 27 March. Looking at the sequence of public health measures in Germany (see e.g. www.acaps.org) and the usual delay between infection and symptoms and reporting (see Linton et al., 2020 and Lauer et al., 2020 for medical evidence on incubation time with median 5.2 days for COVID19 with Chinese data), one could try to identify the events behind these breaks.

In our analysis of the effect of public health measures below, we will focus on RKI data. Hence, we assume that the break took place on 30 March.^4^

### Gompertz curves

The best, almost entirely observation-based, forecast for the evolution of COVID19 in Germany, under the assumption that *RSC do not change*, can be obtained from fitting a Gompertz-curve model to the data. The Gompertz curve is a reduced form, non-linear trend model which is characterized by an upper saturation point which is estimated endogenously. The model displays a double exponential form with three parameters and a time index *t*,

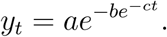

The parameter *b* is a horizontal shift parameter and c is the growth parameter. It can be thought of as the infection rate in this context. The parameter *a* denotes the saturation point: Letting time *t* become larger and larger (we look further and further into the future) shows that *y*_*t*_ approaches *a* as *e*^*-ct*^ with *c* > 0 approaches zero. It is well-known that models of this type capture the s-shape of infection numbers quite well.

Figure 3 summarises the estimated model (employing ordinary least squares and an additive error term). The dark dots are RKI observations and the red dashed curve is the prediction of the model. The green shaded area delineates the 95% confidence region for the forecast. With new data, the green area becomes smaller and approaches the dashed red curve. As this figure impressively shows, Germany seems to be heading towards a stable number of reported COVID19 infections. This number lies at around 184 thousand individuals and would be reached around early May *under the assumption* that current RSC are not modified.

**Figure 3.**
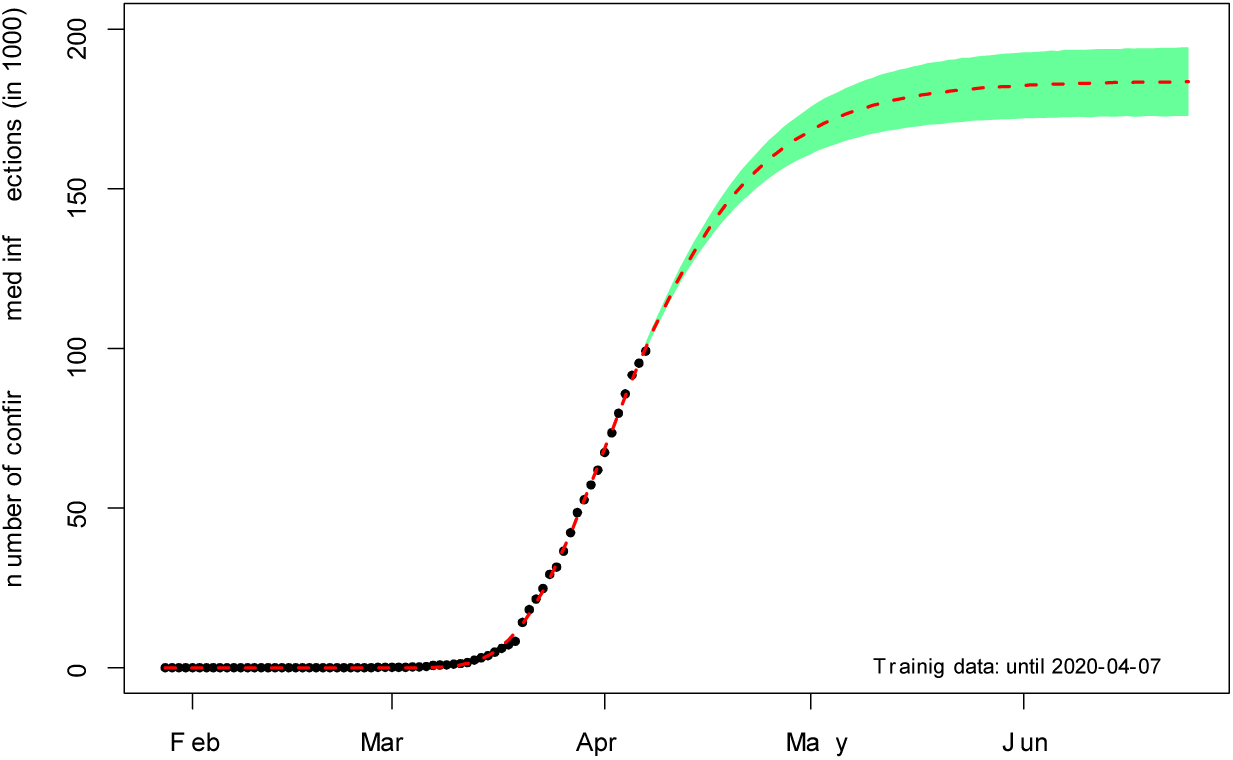
Predicting the number of reported infections (RKI data) under the current regime

## 3 The model

The model was described in detail in Donsimoni et al. (2020). We present only those parts that are important for understanding our calibration below and our forecasts.

### 3.1 The basic structure

The basic structure of the model is illustrated in figure 4. The most well-known background in economics are search and matching models in the Diamond (1982), Mortensen (1982) and Pissarides (1985). The background in mathematics are continuous time Markov chains. We employ this structure and assume four states.

**Figure 4.**
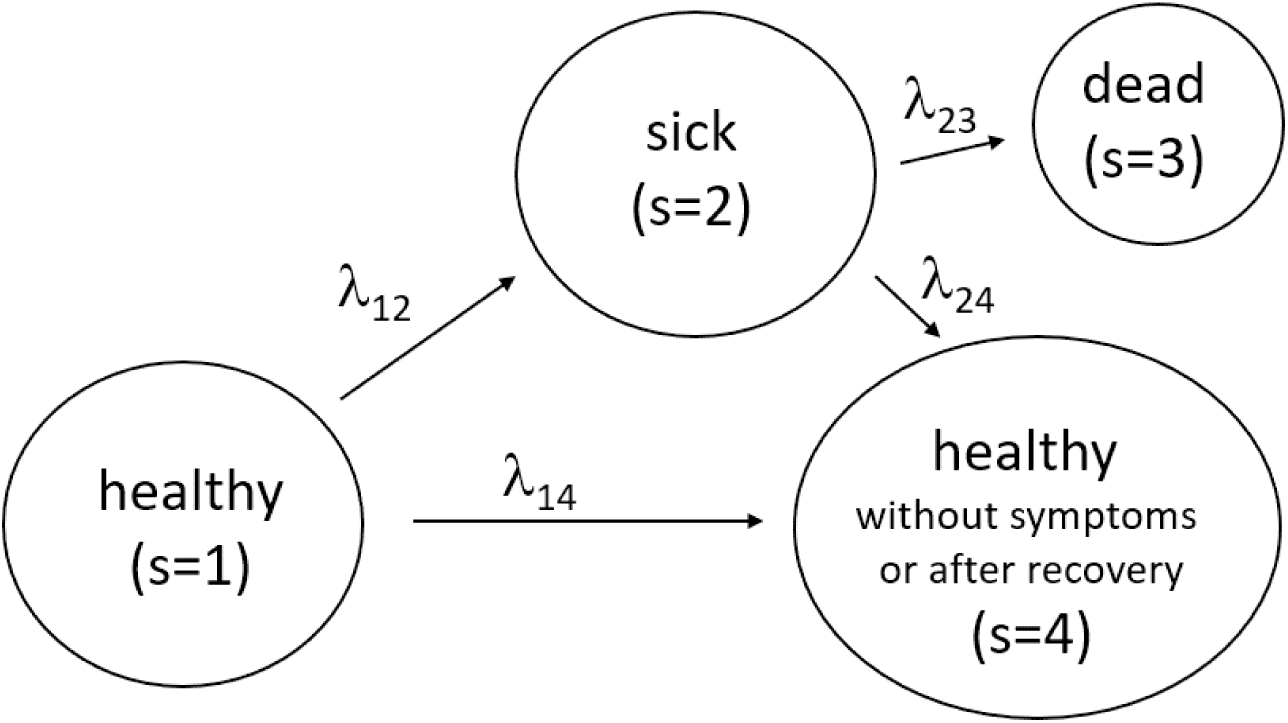
Transitions between the state of health (initial state), sickness, death and health despite infection or after recovery

We employ this figure to offer precise definitions about which individuals we consider to be in which state. State 1 is the state of being healthy in the sense of never having been infected by CoV-2. State 2 captures all individuals that have been *reported* to be infected with SARS-CoV-2. As these reports are based in Germany up to now on tests of individuals that have some (e.g. respiratory) symptoms, we call this the group of sick individuals. The sum of all individuals that are ever reported to be sick is the data collected and published by RKI and JHU that we will employ below. The term sick is also useful as it stresses the differences to individuals that are infected but do not display symptoms. This process is captured in the model by the flows from state 1 to state 4. The size of these flows is a big unknown empirically speaking and several tests are currently being undertaken to measure the number of infected but not sick individuals.^5^ State 3 counts the number of deceased individuals. All individuals that have recovered from being sick or that were never reported or never displayed symptoms after infection are in state 4.

We will employ the terms prevalence and incidence distinctly throughout the paper. Incidence is the number of individuals that are reported for the first time to be sick on a given day. This is the *inflow* into state 2. Prevalence is identical to *N*_2_ (*t*) which denotes the (expected) number of sick individuals at a point in time t in state 2. Prevalence at t is the sum over all incidences from the beginning of the epidemic up to *t* minus the deceased and the recovered individuals.

Data reported by RKI or JHU has traditionally consisted of the number of individuals that were ever reported to be sick, i.e. the sum (integral in terms of the model) of all the inflows into state 2. This quantity at t amounts to prevalence plus the deceased plus the recovered.^6^ The incidence is the daily difference between data reported by RKI or JHU on one day minus the value reported on the day before. This corresponds to incidence above, i.e.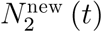 in Donsimoni et al. (2020).

The population in our model is characterized by an infection rate which is simply the ratio of the number of infected individuals (sick and in state 2 or without symptoms in state 4) to individuals that are alive. Letting *N*_*s*_ (*t*) denote the number of individuals in state *s* at *t* the infection rate is simply

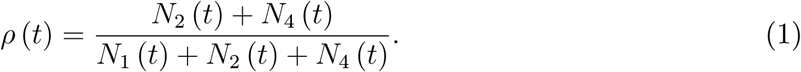

The infection rate is zero initially at *t* < 0. On 24 February 2020 and for Germany, a number of *N*_2_ (0) = 16 sick individuals is introduced into the system and infections and sickness start occurring.

The central transition rate in our model is the individual sickness rate that captures flows from state 1 to state 2. We specify it as

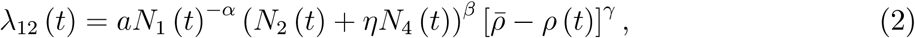

where 0 < α β γ < 1 allows for some non-linearity in the process and *a* > 0. The first term *N*_1_ (*t*)^- α^ captures the idea that more healthy individuals reduce the individual sickness rate. The second term (*N*_2_ (*t*) + 1*N*_4_ (*t*)) ^*β*^ increases the sickness rate when there are more infectious individuals. The parameter 1 describes the fact that individuals that are infected but do not display symptoms (and are therefore in state 4 of our model) nevertheless can infect other individuals. The third term in squared brackets makes sure that the arrival rate is zero when a share 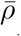-of society is sick (state 2) or healthy after infection (state 4).

The sickness rate satisfies “no sickness without infected individuals”, λ_12_ (*a N*_1_, 0,0. *ρ*) = 0 and “end of spread at sufficiently high level”, 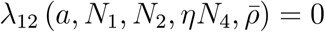. In between these start- and endpoints, the infection rate will first rise and then fall. This specification makes sure that in the long run a share of around 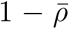 will not have left state 1 i.e. will never have been infected.^7^ We refer to 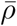 as the long-run share of infected individuals once the epidemic is over.

### 3.2 The model as an ordinary differential equation system

After some steps (see Donsimoni et al., 2020a), our model can be summarized by an ordinary differential equation system. The (expected) number of individuals in state *s* is described by system (3). Parameters not described above are *r* λ_23_ *n*_rec_ and N. The probability to turn sick after an infection with SARS-CoV-2 is denoted by r. The death rate for the transition of sick individuals from state 2 to state 3 visible in figure 4 is denoted by λ _23_. We assume that it takes (on average) *n*_rec_ days to recover from being sick, i.e. to move from state 2 to state 4. Finally, the population size (before the epidemic) is given by *N*.

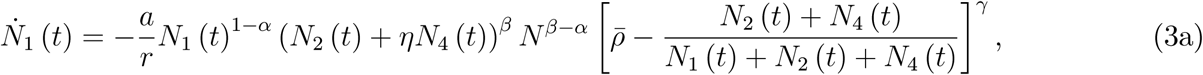

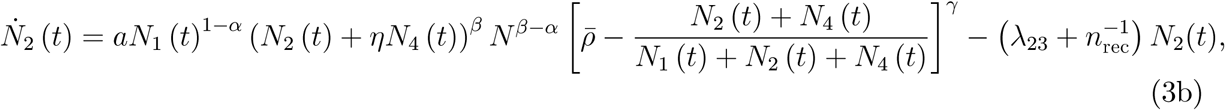

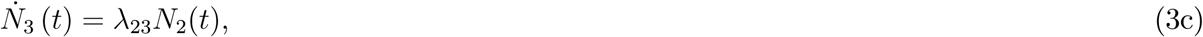

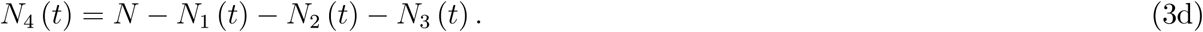

We employ this system for calibration and for prediction. Initial conditions for our solution are 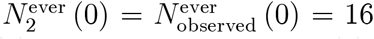 for 24 February 2020 (RKI, 2020), *N*_3_ (0) = *N*_4_ (0) = 0 and 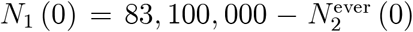, where *N* = 83.1 million is the population size in Germany before the epidemic. Initial conditions for our calibration of the RSC regime are numbers *N*_*s*_ *(t*^*r*^) where *t*^*r*^=30 March 2020 is the day when the RSC regime starts. Initial conditions for predicting the effect of a potential lift of RSC correspond to model predictions for *t*^*l*^=27 April 2020.^8^

## 4 Calibration and model fit

### 4.1 Calibration

The parameters in our model are either chosen exogenously or are the outcome of our data fitting procedure. Exogenous parameters are displayed in table 1.

**Table 1.** Exogenously chosen parameters

| $n_{\text{rec}}$ | $r$ | $\eta$ |
| --- | --- | --- |
| 14 | 0.1 | 0.4 |

As in our earlier work, we assume that recovery takes an average of 14 days. This implies a recovery rate of λ _24_ = 1/14 which captures heterogeneity in the course of the disease (Guan et al., 2020) to some extent. The share *r* of individuals that turns sick (and is reported) after an infection is 10%. The share η of infected individuals without symptoms that can infect other individuals is 40%. See Donsimoni et al. (2020a) for more discussion and robustness analyses.

The model makes a clear prediction about the long-run number of individuals that were ever reported to be sick. As shown in Donsimoni et al. (2020a), this number is given by

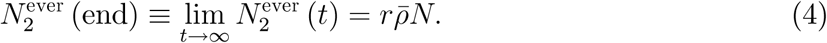

We would like to emphasize that this property of our model is crucial for our long-run predictions and the short-run findings. The long-run number of individuals that, once the epidemic is over, were ever reported to be sick is the probability to get sick after an infection, *r* = 10% times the long-run share of infected individuals, 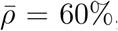 times population size, *N* = 83.1 million, i.e. the long-run number of sick individuals equals 4.99 ≈ 5 million. This is the number of sick individuals in the “normal” scenario of Donsimoni et al. (2020a,b). In their “optimistic Hubei scenario”, they assume that the population share of ever infected individuals once the epidemic is over amounts to 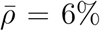 only. In this scenario, the long-run number of sick individuals is 10%× 6% × 83.1 million =498.6 thousand individuals, i.e. roughly 0.5 million individuals. Once this quantity is fixed, any public health measure in our model only shifts the number of sick individuals over the duration of the epidemic. RSC reduces the sickness rate λ _12_ from (2) in the short-run but only delays the infection of the rest of 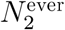 (end) from (4). We admit that this is a strong implication of our model but we only “translate” assumptions made in more general not model-based discussions.^9^

Given that this is a strong assumption and given our Gompertz curve estimation of the current situation in Germany illustrated in figure 3, we are now in the lucky situation that we can do without a strong assumption for 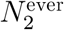 (end) for the *current RSC*. For the current regime (but not for the end of the entire COVID19 epidemic), figure 3 tells us that we are converging in May or June to a value of roughly 184 000 sick individuals. To make clear that this value is valid only for the current RSC, we denote it by 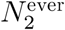 (June) 184 000. This estimate implies a parameter restriction on our long-run value.^10^ Put differently, we can compute

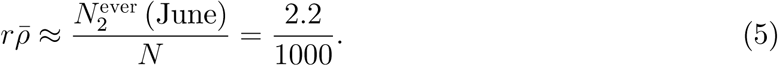

This is the share of sick individuals in the population when the epidemic is over and if the current RSC were preserved forever. The value for the long-run share 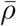 of infected individuals is therefore computed such that (5) is satisfied.^11^

We finally fix various parameters such that we match data reported by RKI. To do so, we minimize the Euclidean distance between the reported data and the predicted values of the model. We undertake two separate calibrations, one for each sub-period described above after the discussion of figure 1. We target a weighted sum of the squared difference between 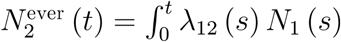 and observation and the newly-sick 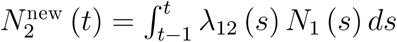 and observation. More precisely, parameters *a*, α, β and γ are obtained from

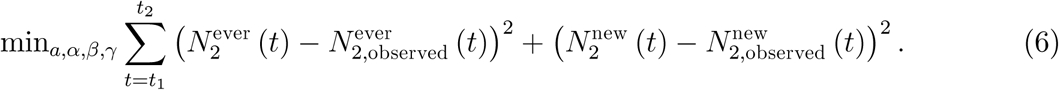

We impose constraints for α, β γ, to lie between zero and one and for *a* to be positive. None of the constraints are binding. Table 2 presents these and all other parameter values both for (*t*_1_, *t*_2_) = (24 Feb to 29 March) and (*t*_1,_ *t*_2_) = (30 March to April).

**Table 2.** Calibrated parameters for RKI data before and after the break

| | $\lambda_{23}$ | $a$ | $\alpha$ | $\beta$ | $\gamma$ | $\bar{\rho}$ |
| --- | --- | --- | --- | --- | --- | --- |
| 24 Feb to 29 March | 1/500 | 3.024/10<sup>6</sup> | 0.5751 | 0.8662 | 0.6459 | 0.06 |
| 30 March to 7 April | 1/500 | 1.48/10<sup>7</sup> | 0.3087 | 0.9511 | 0.7648 | 2.2/100 |

We want to match the number of reported deaths from COVID-19 for our two sub-periods. Hence, the constant death rate for the period from *t*_1_ to *t*_2_ can be computed from

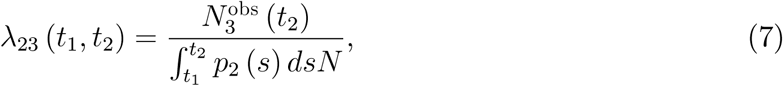

where 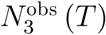 is the number of dead individuals at *T*. Employing this equation yields the values in table 2.

### 4.2 Parameters and model fit

The calibration in Donsimoni et al. (2020a) employed RKI data from 24 February 2020 to *T* = 21 March 2020. Given the impression from figure 1, there is a break in the growth rate of the number of sick only on 30 March (and not on 20 March). We therefore identify two regimes in the RKI data, one from *t*_1_ =24 February to *t*_2_ =29 March and one starting *t*_1_ =30 March.

The calibration results for both regimes are in table 2. The figure also displays 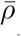 for the pre-RSC regime up to 29 March. We set it equal to 6% and therefore choose the “optimistic Hubei scenario”. The value for 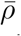 for the RSC regime as of 30 March is the value from (5) divided by *r* from table 1. The death rate λ _23_ is such that the model matches the number of deceased individuals according to (7).

The fit of the calibration can be judged by looking at figure 5.

**Figure 5.**
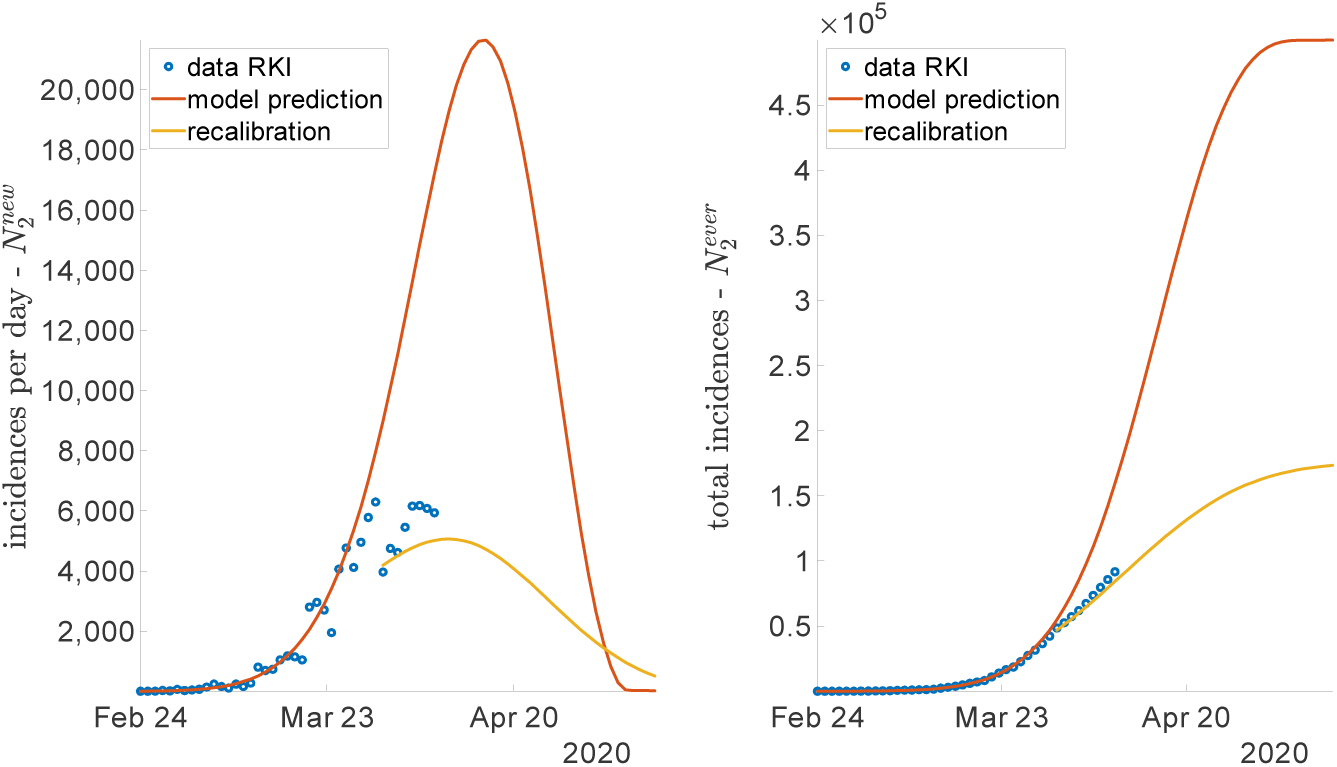
Fit for RKI data, incidences on left and total incidences on right

Our minimization procedure in (6) takes both incidences and total incidences into account without weighting observations explicitly. As a consequence, the fit is unlikely to be equally good. The red curve in the left part of figure 5 shows that incidences up to 29 March are well explained by our model. By contrast, as visible when looking at the yellow curve, daily incidences in the RSC regime are more hard to be captured. We clearly see, however, that the calibrated model already captures the turning point in the number of incidences. This is also what the purely statistical Gompertz approach shown in figure 3 has identified.

The fit for total incidences on the right is very good. The red curve fits data up to 29 March very well and shows where the number reported by RKI would have gone if no RSC had been imposed. The yellow curve shows the numbers one can expect for the weeks to come. From the prediction of the model we are around the turning point now in Germany. The absolute numbers of incidences should now fall on average over the coming weeks. This prediction assumes that public health measures in place do not change *and* that individuals stick to these rules as they used to.

## 5 The effects of RSC and of relaxin them

### The effects of RSC

We now quantify the health effects of restrictions of social contacts (RSC). Our central variable of interest is again the prevalence of COVID19, the number of individuals that are simultaneously sick - *N*_2_ (*t*) in terms of our model. This section also shows what the effects of keeping social distancing forever and relaxing it as of 20 April are.

The blue curve shows the evolution of the epidemic in the absence of any public intervention. This curve employs parameters as calibrated above and as reported in table 1 and the first row of table 2. As the red curve in figure 6 shows, social distancing measures and the shutdown were useful and considerably “flattened the curve”. This curve is plotted using parameter values again from table 1 and from the second row of table 2.

**Figure 6.**
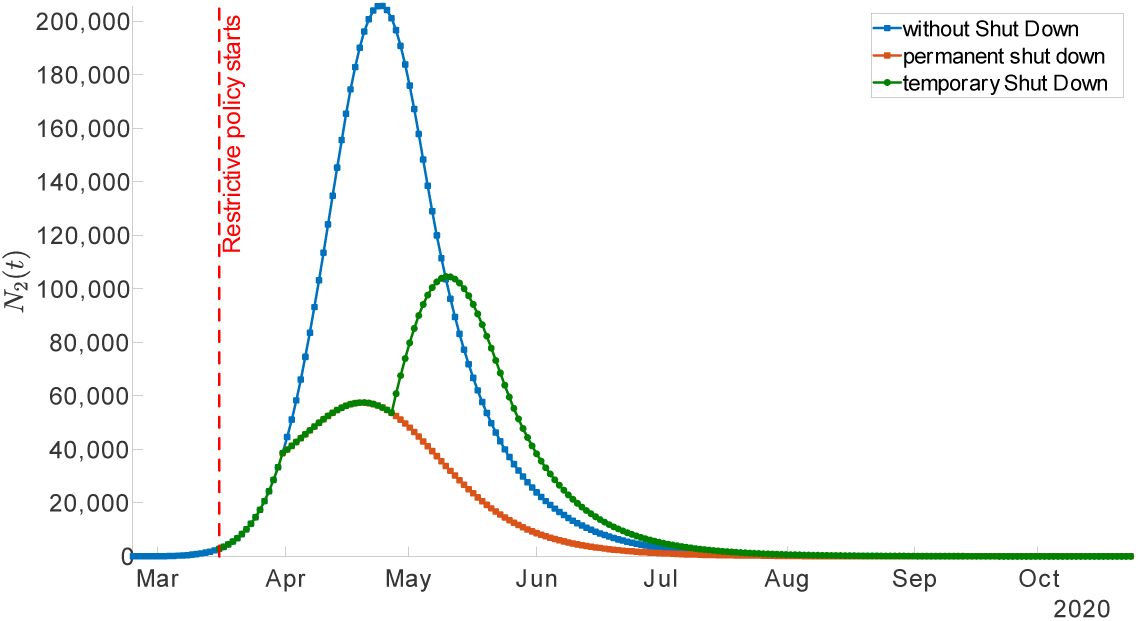
The effect of no shutdown, permanent RSC (shut down) and temporary RSC (shut down) on the prevalence N^2^ (t)

From a pure health perspective this is of course very desirable. As an example, we can again look at the corresponding probabilities to turn sick on a given day or over the period of one week. As the red curve in the left part of figure 5 illustrates, in a situation without RSC, the number of incidences would have continued to increase and so would have the risk to get infected. The yellow curve shows that incidences are now falling and so does the risk to get infected.

While this was expected and predicted by many, our quantitative model can make predictions about the long-run effects of these distancing measures. If measures were *upheld permanently*, the peak of COVID19-prevalence *N*_2_ (t) would be reached end of April already. We can define the end of an epidemic again such that prevalence *N*_2_ (t) falls below 1,000 or the daily incidences are below 100. Prevalence would be lower than 1,000 beginning of July and incidences would be below 100 beginning of May. We stress again that these are expected dates that should hold if RSC are upheld permanently. We also stress that this would not mean a complete end of the epidemic in the sense of herd immunity. There would still be many individuals in state 1 that are not immune and that can be infected and turn sick.

### A complete exit from RSC

Let us return to figure 6 and inquire about the effects of lifting social distancing rules as of 20 April. Due to the delay between infection and reporting also discussed in the context of figures 1 and 2, we assume that the effects of a lift are visible as of 27 April. We therefore plot a green curve in figure 6 that starts on 27 April.

**Figure 2.**
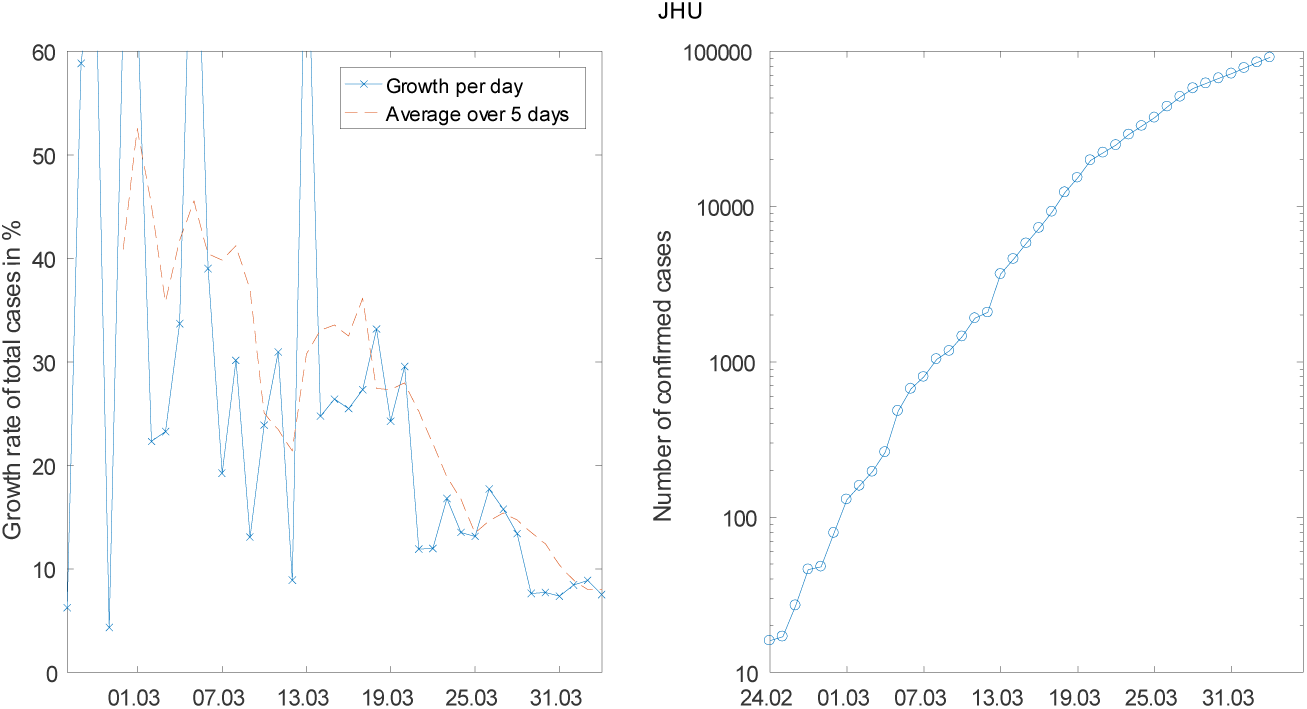
The daily growth rates (left) and the level of the number of sick (right) for JHU data (logarithmic scale)

Plotting this curve requires again parameters for our ODE system in (3). We assume that COVID19 would continue to spread according to the sickness rate λ _12_ from (2). The question is which parameter values we should choose. We do employ parameters in table 1 as always. As it is a projection under a different regime, we cannot employ parameter values from the days before. Hence, concerning parameters from table 2, we assume that the sickness rate is characterized by the same parameter values as before RSC. This leads us to employing the parameter values which we obtained for our calibration of the period from 24 February to 29 March in the first row of table 2.

We should stress that this does not imply that the spread is with the same speed as of 24 February. The number of individuals in states 1 2 and 4, which are the arguments in the sickness rate (2), differ on 27 April as compared to those before any RSC. As a consequence, the speed of the spread will differ.

Plotting the projection for 27 April onwards also requires a value for 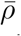. This share of the population that will have been infected once the epidemic is over is the most difficult parameter to be pinned down. If we keep the value of 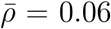^12^ RSC would just imply a shifting of the number of sick over the length of the epidemic. It would, however, not reduce the overall number of sick. It seems natural to assume, however, that RSC not only affect current infection rates but also the long-run share of individuals that are ever infected. We therefore assume a lower value for the long-run infection rate of 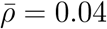.^12^ As is clear from this discussion, a complete lifting of current social distancing rules should lead to an increase in the number of sick individuals again.

Figure 6 therefore summarizes the trade-off decision makers face. Preserving current RSC would be good from a public health perspective but would imply further very high economic costs. A complete lift on 20 April bears the risk of returning to fast growth of the number of sick individuals. The conclusion discusses options that might strike a balance between both scenarios.

## 6 Conclusion

Neither perpetuating the current situation with restrictions on social contacts (RSC) nor a complete lift of RSC is desirable. Preserving the current situation would imply social and economic costs that cannot be sustained for long. Lifting RSC would yield high health risks with a quick increase in the number of sick individuals.

A way out must consist in measures that reduce economic costs without increasing infection risks substantially (see Abele-Brehm et al., 2020, for suggestions). At the same time one should not follow a one-rule-fits-all policy for all regions in Germany. If different regions (or even smaller communities) run different policies and data is well-recorded for smaller areas as well, decision makers could quickly learn about which measures are most effective in terms of reducing infection rates as well as reducing economic and social costs. As an example, some regions could allow for schools to open again as of grade 9, others only as of grade 5. Other regions could allow restaurants (preserving a distance of 2 meters between tables) to open, while others do not. A trial period of four weeks with partially relaxed rules in some parts of Germany should be enough to identify the effects. One should then be prepared to adjust the measures (both upwards or downwards depending on the outcomes) in around four weeks after relaxing the measures.

These measures would not be required if truly large-scale testing of the population and isolation of infected and sick was possible. In the absence of medical testing, one can only learn by coordination of heterogeneous regional responses to COVID19. This would be a good example of how a federal system can be used to learn from each other. If this option is ignored, it will be just as difficult in one month’s time to judge which measures help economically and are not too costly from a health perspective.

## Data Availability

Data used is publicly available.

https://raw.githubusercontent.com/CSSEGISandData/COVID-19/master/csse_covid_19_data/csse_covid_19_time_series/time_series_covid19_confirmed_global.csv

## Footnotes

2 See Donsimoni et al. (2020b) for a summary in German.

3 These three papers were presented at the ‘Forecasting COVID19’ workshop at the Johannes Gutenberg University on 6 April 2020.

4 We have undertaken analyses with JHU data as well where we assumed that the effects of public health measures are visible as of 20 March. While there are obviously (small) quantitative differences, the broad picture remains the same.

5 Our earlier paper (Donsimoni et al. 2020) discusses in detail how we quantify this flow. The crucial assumption concerns the share of infected individuals that do not display symptoms or are not reported. We assume this share is around 80% to 90%. In terms of model parameters, this means we assume *r* = 10% (see below).

6 We denote this by 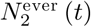 in Donsimoni et al. (2020).

7 We employ “around” as some individuals will have ended up in state 3 whose number does not enter the expression in (1).

8 As discussed below, we assume that a lift on 20 April would imply observable effects only around one week later.

9 In ongoing work we study the historical evidence about 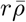 from other epidemics and pandemics. No systematic evidence seems to be available at this point. We are grateful to dozens of epidemiologists, virologists, economists and decision-makers for discussions of this point.

10 We are grateful to Hilmar Schneider for having raised this point.

11 We emphasize again that preserving the RSC would be unlikely to set an end to the epidemic as some infected individuals will remain within the population also by June. A lift of RSC only in June would then lead to a next rise of infections.

12 We emphasize that this is the outcome of many comments and discussions about the effects of shutdowns on long-run infection rates. It is generally argued that more social separation does not only reduce the infection rate instantaneously but also in the long run.

## References

Abele-Brehm, A., H. Dreier, C. Fuest, V. Grimm, H.-G. Krausslich, G. Krause, M. Leonhard, A. Lohse, M. Lohse, T. Mansky, A. Peichl, R. M. Schmid, G. Wess, and C. Woopen (2020): “Die Bekampfung der Coronavirus-Pandemie tragfahig gestal-ten,” https://www.ifo.de/publikationen/2020/monographie-autorenschaft/die-bekaempfung-der-coronavirus-pandemie-tragfaehig.

Adamik, B., M. Bawiec, V. Bezborodov, W. Bock, M. Bodych, J. Burgard, T. Gatz, T. Krueger, A. Migalska, B. Pabjan, T. Ozanski, E. Rafajlowicz, W. Rafajlowicz, E. Skubalska-Rafajlowiczc, S. Ryfczynska, E. Szczureki, and P. Szymanski (2020): “Mitigation and herd immunity strategy for COVID-19 is likely to fail,” Working Paper TU Kaiserslautern.

Dehning, J., J. Zierenberg, F. P. Spitzner, M. Wibral, J. P. Neto, M. Wilczek, and V. Priesemann (2020): “Inferring COVID-19 spreading rates and potential change points for case number forecasts,” mimeo Max Planck Institute for Dynamics and Self-Organization, Gattingen.

Diamond, P. A. (1982): “Aggregate Demand Management in Search Equilibrium,” Journal of Political Economy, 90, 881–94.

Donsimoni, J. R., R. Glawion, B. Plachter, and K. Waelde (2020): “Projektion der COVID-19-Epidemie in Deutschland,” Wirtschaftsdienst, https://www.wirtschaftsdienst.eu/inhalt/jahr/2020/heft/4/beitrag/projektion-der-covid-19-epidemie-in-deutschland.html, 100(4), 247–249.

Donsimoni, J. R., R. Glawion, B. Plachter, and K. Walde (2020): “Projecting the Spread of COVID19 for Germany,” revise & resubmit, https://www.medrxiv.org/content/10.1101/2020.03.26.20044214v1.

Gros, C., R. Valenti, K. Valenti, and D. Gros (2020): “Strategies for controlling the medical and socio-economic costs of the Corona pandemic,” https://arxiv.org/abs/2004.00493.

Guan, W., Z. Ni, Y. Hu, W. Liang, C. Ou, J. He, L. Liu, H. Shan, C. Lei, d. Hui, B. Du, L. Li, G. Zeng, K.-Y. Yuen, R. Chen, C. Tang, T. Wang, P. Chen, J. Xiang, S. Li, J.-l. Wang, Z. Liang, Y. Peng, L. Wei, Y.-h. Hu, P. Peng, J.-m. Wang, J. Liu, Z. Chen, Z. Li, Z. Zheng, S. Qiu, J. Luo, C. Ye, S. Zhu, and N. Zhong (2020): “Clinical Characteristics of Coronavirus Disease 2019 in China,” The New England Journal of Medicine, doi: https://doi.org/10.1101/2020.02.06.20020974, p1-12.

Hartl, T., K. Walde, and E. Weber (2020): “Measuring the impact of the German public shutdown on the spread of COVID19,” Covid economics, Vetted and real-time papers, CEPR press, 1, 25–32.

Lauer, S., K. Grantz, Q. Bi, F. Jones, Q. Zheng, H. Meredith, A. Azman, N. Reich, and J. Lessler (2020): “The Incubation Period of Coronavirus Disease 2019 (COVID-19) From Publicly Reported Con?rmed Cases: Estimation and Application,” Annals of Internal Medicine, doi:10.7326/M20-0504, 1–7.

Linton, N., T. Kobayashi, Y. Yang, K. Hayashi, A. R. Akhmetzhanov, S.-m. Jung, B. Yuan, R. Kinoshita, and H. Nishiura (2020): “Incubation Period and Other Epidemiological Characteristics of 2019 Novel Coronavirus Infections with Right Truncation: A Statistical Analysis of Publicly Available Case Data,” Journal of Clinical Medicine, 9(2)(538), 1–9.

Johns Hopkins University (2020): “COVID19 dataset,” https://raw.githubusercontent.com/CSSEGISandData/COVID-19/master/csse_covid_19_data/csse_covid_19_time_series/time_series_covid19_confirmed_global.csv.

Mortensen, D. T. (1982): “Property Rights and Efficiency in Mating, Racing, and Related Games,” American Economic Review, 72, 968–79.

Pissarides, C. A. (1985): “Short-run Equilibrium Dynamics of Unemployment Vacancies, and Real Wages,” American Economic Review, 75, 676–90.

Robert Koch Institut (RKI) (2020): “COVID-19: Fallzahlen in Deutschland und weltweit,” https://www.rki.de/DE/Content/InfAZ/N/Neuartiges_Coronavirus/Fallzahlen.html.

